# Performance Optimization of an R Shiny-based Digital Health Dashboard for Monitoring Small and Sick Newborn Care in Low-resource Hospital Settings

**DOI:** 10.64898/2026.03.08.26347893

**Authors:** Julius Thomas, Georgia Jenkins, Junwei Chen, Morris Ogero, Lucas Malla, Lisa R Hirschhorn, Rebecca Richards-Kortum, Z. Maria Oden, Christine Bohne, John Wainaina

**Affiliations:** Rice360 Institute for Global Health Technologies, Rice University, Texas, USA; London School of Hygiene and Tropical Medicine, London, United Kingdom; Department of Medical Social Science, Feinberg School of Medicine, Northwestern University, Chicago, USA

**Keywords:** R Shiny, dashboard, neonatal, optimization, low-resource settings, hospitals, data

## Abstract

**Background:** Digital health dashboards can enhance health system performance by transforming routinely collected data into actionable insights for decision-making. In low-resource settings, however, their effectiveness depends not only on the relevance of indicators but also on system reliability within constrained digital infrastructure. Neonatal mortality remains a major global health challenge, with the highest burden in low- and middle-income countries, where many deaths are preventable through timely, evidence-based interventions. Continuous monitoring of care processes and outcomes is therefore essential. To support this need, we developed the NEST360 Implementation Tracker (NEST-IT) using R Shiny to support quality improvement across more than 100 hospitals in sub-Saharan Africa. As the platform scaled to over half a million records and increasing concurrent users, performance constraints emerged, particularly in hospitals with limited computing resources, threatening timely access to critical information.

**Objective:** This study aimed to describe optimization strategies applied to the NEST-IT dashboard and evaluate their impact before and after implementation.

**Methods:** A structured optimization process was implemented following established R Shiny performance principles. Dashboard profiling was first conducted to identify key bottlenecks, after which targeted improvements were applied to improve efficiency and responsiveness. A quasi-experimental pre–post evaluation (December 2023–August 2024) assessed performance using three indicators: server processing time, visualization rendering time (VRT), and Time to First Byte (TTFB). Metrics were measured repeatedly during one-month baseline and post-optimization periods and summarized using mean values.

**Results:** Four primary bottlenecks were identified: delayed server responses, slow visualization rendering, inefficient data handling, and inconsistent device performance. Following optimization, interactive plot load time decreased from 10.1 to 2.7 ± 0.6 seconds (73.3% improvement). Visualization rendering improved from 3.61 to 1.62 seconds, while server processing time fell from 2.3 ± 0.7 to 0.8 ± 0.3 seconds. TTFB improved from 1.9 ± 0.4 to 0.6 ± 0.2 seconds, and system uptime increased from 92.5% to 99.2%.

**Conclusion:** Performance optimization substantially improved dashboard responsiveness, enabling timely access to critical neonatal information in resource-constrained hospital settings. The findings provide practical, evidence-based framework for improving the performance of R Shiny dashboards and demonstrate scalable strategies for delivering reliable digital decision-support tools in low-resource health systems.

## 1 Introduction

Digital health dashboards are increasingly used to strengthen health system performance by translating routinely collected data into actionable insights for clinicians, managers, and policymakers (1, 2). In low-resource hospital settings, the effectiveness of such dashboards depends not only on the relevance of the indicators displayed, but also on system performance, responsiveness, and usability under constrained infrastructure (3, 4). Optimized dashboards can enable continuous monitoring of key indicators, support timely decision-making, and facilitate targeted quality improvement interventions in low- and middle-income countries (LMICs) (1, 2).

Small and sick newborn care provides a compelling use case for dashboard-supported decision-making. Neonatal deaths account for a substantial proportion of under-five mortality globally, with the greatest burden in LMICs (5, 6). Many of these deaths are preventable through a comprehensive strategy that combines routine monitoring of critical neonatal indicators (7–9), and the timely implementation of evidence-based interventions, including Kangaroo Mother Care (KMC) (10), thermal care, infection prevention and control measures, and appropriate respiratory support using Continuous Positive Airway Pressure (CPAP) or oxygen therapy (11).

Regular monitoring of key indicators in neonatal care is critical for identifying gaps in service delivery. Early identification of these gaps enables timely adjustments and targeted implementation of interventions at the facility level and key decision-making at national level including building data use capacity and quality improvement (9, 12). To support such continuous monitoring and enable timely, evidence-informed decisions, interactive data visualization tools such as dashboards have become increasingly valuable. These tools simplify complex clinical data into clear, actionable insights, enabling health care providers and managers to monitor care quality and respond promptly to emerging challenges (13). This is especially important in low-resource hospital settings, where small and sick newborns face a significantly higher risk of complications and death, and where timely guidance and tracking of care practices are critical (14).

To meet this need, we developed the NEST360 Implementation Tracker (NEST-IT) dashboard, an interactive data platform designed to support monitoring of essential neonatal care interventions and contextual factors. Built using the R Shiny framework, an open-source web application framework for R that enables the creation of interactive data visualization tools, the dashboard integrates diverse data sources into a single, user-friendly interface tailored for clinicians, hospital leadership, national Ministry of Health leadership, program managers, funders and other relevant stakeholders to perform intuitive monitoring and visualization of neonatal care. By improving access to timely and actionable data, the NEST-IT dashboard also enables the NEST360 team to make swift, informed, data-driven decisions ultimately improving neonatal healthcare delivery in low-resource settings. The NEST360 Alliance currently spans multiple countries across sub-Saharan Africa, including Malawi, Tanzania, Kenya, Nigeria and Ethiopia, supporting implementation in more than 100 hospitals, thereby providing a robust regional platform for advancing small and sick newborn care at scale (15).

However, as the platform scaled to support over half a million records, manage more than 250 concurrent users and 8 distinct user groups and expand hospital coverage from 68 to over 100 facilities, the limitations of R Shiny’s single-threaded architecture became increasingly evident (16, 17). These constraints manifested as longer page load times, reduced responsiveness of interactive components, and significant delays in rendering complex visualizations, ultimately impacting the user experience and operational efficiency of the dashboard.

These performance challenges are even more significant in environments characterized by constrained computational resources, for example, settings where hospitals and clinics have desktop computers with outdated hardware, limited internet connectivity, and low processing power, or difficulties in use due to incompatible devices such as personal phones and tablets (13). In such conditions, any decrease in dashboard speed or functionality directly impacts its usability and can delay critical clinical decision making (18, 19).

Although R Shiny is increasingly adopted in global health settings, largely because it is open-source, cost-effective, supports rapid prototyping, and enables the development of full-featured web applications (20), there remains a shortage of practical, context-specific guidance on how to optimize its performance under resource-constrained conditions. This paper aims to address that gap by detailing the optimization strategies applied to enhance the performance of the NEST-IT dashboard. Specifically, we identify and describe key techniques including asynchronous processing, reactive and vectorized programming practices, caching mechanisms, database query optimization, and load balancing across servers.

We also evaluate how these strategies improved dashboard performance by comparing performance before and after the implementation of the targeted optimization techniques. By documenting these enhancements, we aim to offer practical, evidence-based recommendations for developers and health system implementers seeking to build high-performing, scalable dashboards suited to the infrastructural constraints of low-resource healthcare settings.

## 2 Materials and methods

### 2.1 Study Design

A quasi-experimental pre-post study design was conducted between December 2023 and August 2024 to evaluate changes in dashboard performance following the implementation of a series of optimization strategies in the NEST-IT dashboard.

### 2.2 Stepwise Optimization Framework

Dashboard optimization followed a structured five-phase framework consisting of performance profiling, bottleneck identification, strategy selection, implementation, and evaluation (Figure 1).

**Figure 1.**
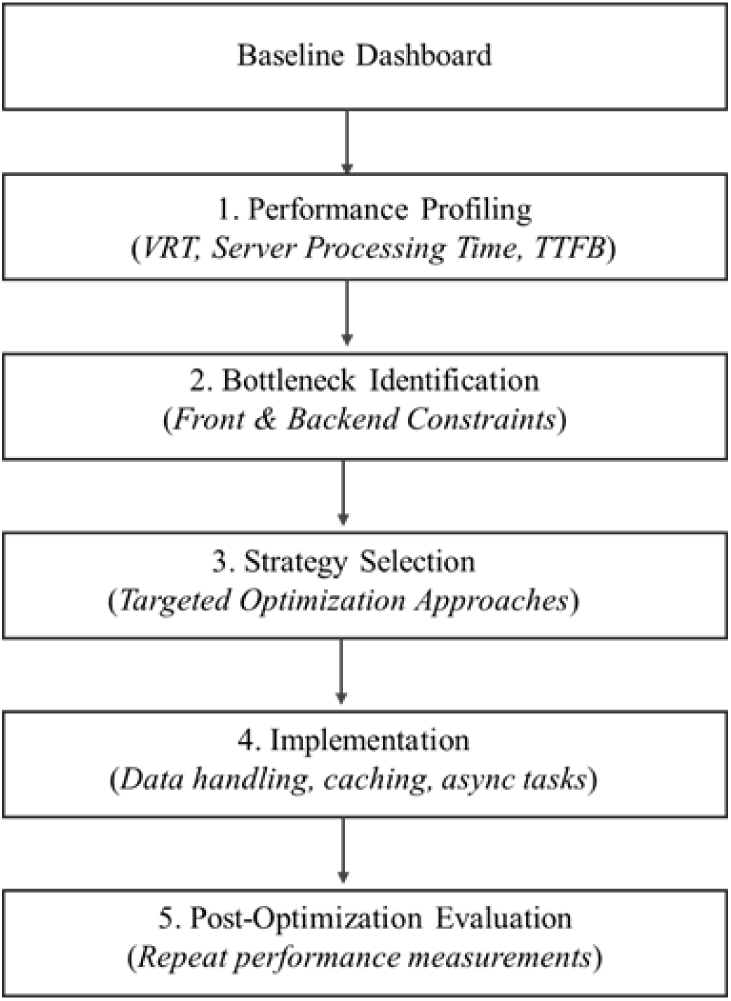
The NEST-IT Dashboard Optimization Framework Workflow

#### 2.2.1 Performance Profiling

Baseline performance was assessed using three key metrics capturing both frontend and backend responsiveness (Table 1).

**Table 1.** Dashboard Performance Metrics Used for Profiling.

| Metric | Description | Measurement Method | Bottleneck Threshold |
| --- | --- | --- | --- |
| Visualization Rendering Time (VRT) | Time from visualization request to full rendering in the interface (load time) | Browser developer tools | > 4 seconds |
| Server Request Processing Time | Time required for the server to process filtering or computation tasks | profvis profiling and R timing logs | > 2 seconds |
| Time to First Byte (TTFB) | Time between request initiation and receipt of the first byte of data | Network timing tools across multiple devices | > 1 second |

Each metric was measured five times per visualization, and mean values were used for analysis. Load time benchmarks were aligned with established usability standards for interactive systems (21, 22). Visualizations with average rendering times exceeding 4 seconds were classified as unresponsive and flagged for optimization. Similarly, dashboard components with server processing times above 2 seconds were considered backend performance bottlenecks. Consistent with established performance benchmarks (23, 24), dashboard sections with average TTFB values exceeding 1 second were prioritized for optimization.

#### 2.2.2 Bottleneck Identification

Sections exceeding the predefined thresholds were flagged as performance bottlenecks, and their causes classified accordingly (Table 2).

**Table 2.** Common Performance Bottlenecks in NEST-IT Dashboard.

| Bottleneck Category | Underlying Causes |
| --- | --- |
| Slow visualization rendering | Processing of large datasets during plot generation, inefficient data manipulation routines, and computationally intensive indicator calculations performed at runtime |
| Delayed server responses | Repeated re-computation of reactive objects and inefficient dependency management within the dashboard's reactive framework |
| Inefficient data pipelines | Sequential file loading, redundant data transformations, and direct ingestion of large raw datasets without prior preprocessing or caching |
| Inconsistent device performance | Non-optimized cross-platform layout rendering and varying computational capacities across devices, particularly low-powered smartphones and tablets |

This process generated a systematic mapping of performance constraints that informed the selection of optimization strategies.

#### 2.2.3 Strategy Selection

Optimization strategies were selected based on the bottlenecks identified during the profiling stage. Guided by established R Shiny optimization practices reported in previous studies (25, 26), we prioritized strategies that directly addressed the specific constraints identified in the bottleneck analysis.

#### 2.2.4 Implementation of Selected Optimization Strategies

A suite of optimization strategies was implemented iteratively to enhance the responsiveness, scalability, and reliability of the NEST-IT dashboard (Table 3). Each strategy targeted specific performance constraints identified during the bottleneck profiling phase, ensuring that the platform can consistently support the NEST360 program by delivering timely, data-driven insights for decision-making.

**Table 3.** Optimization Strategies Implemented.

| Optimization Strategy | Specific Implementation Technique | Purpose |
| --- | --- | --- |
| Efficient Data Handling | Offline preprocessing | Raw data is cleaned, filtered, and aggregated offline before being loaded into the dashboard, reducing runtime computation |
|  | Lazy loading | Only the variables required for a specific task are loaded into memory from the raw dataset (Figure S1), This reduces memory consumption and load times |
|  | Parallelized bulk data loading | Accelerate multi-file ingestion (Figure S2). This reduces initialization time for large, multi-file datasets |
| Reactive Programming | Real-time Updates | Ensure plots and tables update automatically only when their inputs change. (Figure S3A & S3B). This eliminates redundant recalculations |
|  | Integration with memoization | Combining *reactive()* with *memoise()* caches result for specific input combinations, so returning to a previous view is instantaneous (Figure S4) |
| Caching | Memoization | Cache results of complex computations to avoid redundant processing (Figure S5) |
| Asynchronous Processing | *future()* and *promises* | Long-running computations are executed in a separate process, keeping the main Shiny session responsive and preventing UI freezes (Figure S6) |
| Mobile-friendly Layout | JavaScript-enabled Layout | Responsive design adaptations implemented in JavaScript ensure usability and efficiency on low-powered mobile devices and tablets (Figure S7A & S7B) |
| Vectorized Programming | *dplyr* / *data.table* vectorization | Replace loops with optimized operations, drastically improving performance on large datasets (Figure S8) |

Performance improvements were quantified by comparing pre- and post-implementation delay times across key performance metrics. Representative code snippets illustrating these optimization approaches are provided in the Supplementary Materials to enhance clarity, reproducibility, and practical application.

#### 2.2.5 Post-Optimization Evaluation

Following implementation of the optimization strategies, dashboard performance was reassessed using the same metrics applied during baseline profiling, enabling direct comparison of frontend and backend responsiveness before and after optimization (Table 4).

**Table 4.** Post-Optimization Evaluation Framework.

| Metric | Purpose |
| --- | --- |
| Visualization Rendering Time (VRT) | Assess frontend rendering improvements |
| Server Request Processing Time | Evaluate backend computation efficiency |
| Time to First Byte (TTFB) | Measure overall server responsiveness |

Each metric was recorded five times per visualization across multiple sessions and device types. Mean values were calculated to account for variability in real-world usage conditions and to provide representative estimates of dashboard performance.

### 2.3 Comparison of Baseline and Post-Optimization Performance

Performance outcomes were systematically compared between baseline and post-optimization assessments to determine the extent of improvement following optimization. Visualization load times, server-side processing durations and TTFB were documented both before and after intervention, with mean values derived across five repeated trials. Continuous monitoring of all three metrics was carried out iteratively as optimization strategies were introduced, and the process was sustained until predefined performance thresholds were achieved (VRT < 4 seconds, Server Request Processing Time < 2 seconds, and TTFB < 1 second).

Paired t-tests were used to compare baseline and post-optimization values for visualization load time, server-side processing time, and TTFB. These tests were selected because the outcomes are continuous measures, and preliminary inspection suggested approximate normality of paired differences. Given the modest number of repeated trials, robustness was assessed using the non-parametric Wilcoxon signed-rank test, which does not assume normality.

Data were systematically recorded by dashboard developers during iterative testing and analyzed to assess the degree of performance gains. This comparative approach provided a rigorous evaluation of the optimization strategies, enabling us to establish both the magnitude and consistency of improvements in system responsiveness across different dashboard sections.

## 3 Results

### 3.1 Baseline Performance Bottlenecks

Baseline performance profiling of the NEST-IT dashboard revealed four key bottlenecks affecting system responsiveness across both backend computation and frontend visualization processes. These constraints were identified through a combination of quantitative performance measurements (Table 5) and qualitative usability observations during cross-device testing (Table 6).

**Table 5.** Quantitatively Measured Performance Bottlenecks Identified During Profiling.

| Bottleneck Category | Baseline Observation | Baseline Measurement |
| --- | --- | --- |
| Slow visualization rendering | Repeated loading of unchanged data | > 4 seconds |
|  | Prolonged app initialization and login delays |  |
|  | Slow loading time of plots |  |
| Delayed server responses | Heavy data processing increased backend processing time | 2.3 ± 0.7 seconds |
| Inefficient data handling | Loading large raw datasets increased dashboard startup time | 9 seconds |

**Table 6.** Performance Bottlenecks Identified Through Device Usability Testing.

| Bottleneck Category | Baseline Observation | Identification Method |
| --- | --- | --- |
| Inconsistent device performance | Reduced responsiveness and distorted layout during low-powered mobile devices usage | Cross-device usability testing on smartphones and tablets |

### 3.2 Optimization Strategies Implemented

To systematically address these challenges, six targeted optimization strategies were developed and implemented, guided by best practices and evidence from the literature. Each strategy targeted a specific performance bottleneck and was applied stepwise. Table 7 summarizes the correspondence between the identified issues, the optimization measures undertaken, the expected performance and improvements in system performance.

**Table 7.** Optimization Strategies Performance.

| Optimization Strategy | Bottleneck Addressed | Expected Performance Gains | Time Improvement (95% CI) |
| --- | --- | --- | --- |
| Efficient data handling | Inefficient processing of large datasets | Faster dashboard start-up time, and faster UI rendering | 40% (34–46) decrease in responsiveness |
| Lazy Loading | Slow loading time and memory overload | Faster loading; reduced memory usage | 51% (41–56) decrease in time-to-first-interaction |
| Reactive Programming | Untimely data visualization | Near real-time interactivity and responsiveness | 94% (90–98) reduction in time |
| Caching Mechanisms | Repeated loading of unchanged data | Faster data retrieval; improved dashboard load speed | 53% (44–61) reduction in load time |
| Asynchronous processing | Prolonged app initialization and login delays | Faster login and plot loading | 45% (38–56) reduction in initialization time |
| JavaScript Interface Adaptability | Incompatible device support | Optimized layout for low-powered devices (phones and tablet) | 32% (26–30) reduction in device-specific lag |
| Vectorized Programming enhancements | Slow computations and data transformations | Faster execution of data workflows; smoother user interactivity | 48% (39–55) reduction in computation time |

Optimizations targeting key bottlenecks, particularly Reactive Programming, Caching, and Lazy Loading, substantially improved dashboard performance, with complementary strategies further enhancing computation, UI responsiveness, and accessibility on low-powered devices.

### 3.3 Pre- vs Post-Optimization Performance

Comparison of baseline and post-optimization performance metrics was conducted using both parametric (paired t-test) and non-parametric (Wilcoxon signed-rank) approaches. Paired t-tests indicated statistically significant reductions across all performance metrics (p < 0.05). Corresponding Wilcoxon signed-rank tests produced consistent results, supporting the robustness of these findings (Table 8).

**Table 8.** Baseline vs Post-Optimization Performance.

| Metric | Mean (Baseline) | Mean (Post) | Mean Diff. (95% CI) | Paired t-test (p) | Wilcoxon test (p) |
| --- | --- | --- | --- | --- | --- |
| Visualization Load Time (s) | 3.62 | 1.62 | -2.0 (-2.55, -1.45) | <0.001 | 0.058 |
| Server Processing Time (s) | 2.30 | 0.80 | -1.5 (-2.05, -0.96) | <0.001 | 0.063 |
| Time to First Byte (TTFB) (s) | 1.90 | 0.60 | -1.3 (-1.62, -0.98) | <0.001 | 0.062 |

Optimization produced substantial improvements across all performance metrics (Table 8). Visualization load time decreased by 2.0 s (95% CI: -2.55, -1.45), server processing time by 1.5 s (95% CI: -2.05, -0.96), and TTFB by 1.3 s (95% CI: -1.62, -0.98). Paired t-tests confirmed statistical significance (p < 0.001), and Wilcoxon signed-rank tests produced consistent results, supporting robustness given the modest sample size.

### 3.4 Visualization Load Times of Complex Plots

Tables Optimization strategies greatly enhanced the performance and responsiveness of the NEST-IT dashboard, significantly reducing loading times across all visualization components. The delays in processing and rendering the complex plots are detailed in Figure 2.

**Figure 2.**
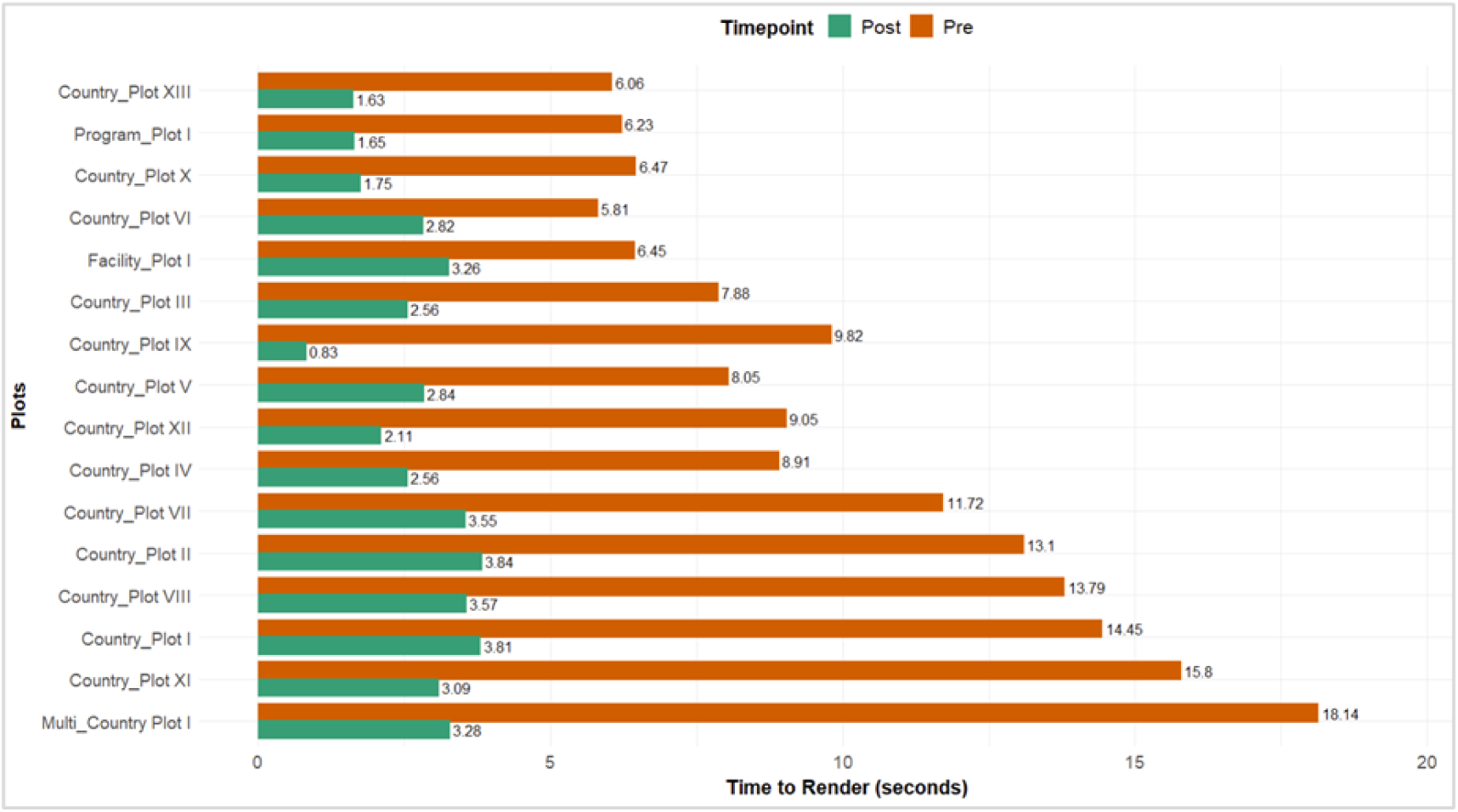
Time to render Complex Plots Pre and Post Optimization

Prior to optimization, complex visualizations, such as interactive plots, took an average of 10.11 ± 1.1 seconds to render, while tables and static content experienced moderate delays of 1.8 ± 1.4 seconds. More specifically, longer delays in rendering was observed in complex plots which had multiple countries or multiple facilities to process and render, some including multi-country plot I:18.14s, country plot XI: 15.8s, country plot I:14.45s. Such delays are well above accepted usability thresholds and would likely disrupt user workflow.

Post-optimization, interactive plots, previously the most affected by performance bottlenecks saw a 73.3% reduction in average loading time, rendering in 2.7 ± 0.6 seconds. Overall, all plots loaded in 1.62 ± 0.4 seconds, down from 3.61 ± 0.4 seconds, reflecting a 2.2-fold improvement, server processing time fell from 2.3 ± 0.7 to 0.8 ± 0.3 seconds, while TTFB improved from 1.9 ± 0.4 to 0.6 ± 0.2 seconds. The most pronounced improvements were observed in multi-country plot I (reduced from 18.14s to 3.28s, ∼82% reduction), country plot XI (15.8s to 3.09s, ∼80% reduction), and country plot I (14.45s to 3.81s, ∼74% reduction).

Successive rounds of performance tuning ensured that all visualization components met the predefined performance threshold (<4 seconds), enhancing real-time usability for healthcare providers. Additionally, system stability improved, with the dashboard maintaining 99.2% uptime post-optimization, compared to 92.5% at baseline, mitigating failures caused by high data loads and increased user sessions and use. These optimizations collectively improved system throughput, ensuring that dashboard users experienced minimal latency and faster data retrieval.

## 4 Discussion

The performance optimization of the NEST-IT dashboard demonstrates that carefully selected architectural and computational strategies can substantially improve the responsiveness of R Shiny–based applications in low-resource hospital settings. Collectively, these optimizations directly addressed the inherent limitations of R Shiny’s single-threaded architecture (16, 17), reducing computational bottlenecks and ensuring reliable performance under real-world conditions characterized by high data volumes and diverse device environments. As a result, rendering times for complex plots were reduced by up to 82%, with all visualizations consistently loading in under four seconds, a threshold commonly cited as critical for maintaining user engagement in interactive dashboards (21, 22, 27). By directly reducing prolonged response delays, these improvements directly address the usability barrier of prolonged delays in data-intensive dashboards and establish an optimized, scalable architecture suitable for health programs in resource-constrained contexts.

Prior to optimization, high data loads and complex visualizations resulted in slow response times, limiting timely decision-making in low-resource hospital settings. Of the strategies implemented, reactive programming emerged as the most impactful, achieving approximately 94% improvement in rendering times and enabling near-instant interactivity for visualizations. This advancement ensured that stakeholders could access and interpret information without delay, thereby supporting rapid insights and life-saving decisions for vulnerable newborn populations. These findings are consistent with earlier reports that underscore the importance of reactive programming frameworks in building scalable, real-time applications. For instance, a performance evaluation conducted in China demonstrated that server-side reactive programming excels under data-heavy, real-time web workloads, maintaining low latency and stable throughput even at high levels of concurrency by avoiding blocking operations and inefficient thread utilization (28). By synchronizing user actions with immediate updates, reactive programming can help reduce the lag experienced during dashboard use and may reinforce the clinical value of dashboards as dependable, time-sensitive decision-support tools.

In addition to reactive execution, the integration of caching mechanisms played a pivotal role in improving responsiveness. By minimizing redundant data retrieval and reuse of previously computed results, caching reduced load times by more than half. Similar approaches in clinical dashboards have been shown to significantly improve user experience and system scalability (26, 29). For instance, the SAS BI Dashboard employs an in-memory Least Recently Used (LRU) caching framework that serves dashboard data directly from cache when the underlying data remain current, refreshes stale content on demand, and uses background processes to proactively update cached models, thereby minimizing repeated database queries and accelerating dashboard rendering (30). In NEST-IT, caching ensured that repeated queries to unchanged datasets no longer imposed delays, a feature particularly useful in environments where healthcare providers frequently revisit the same metrics during patient monitoring or program evaluations, with minimal latency.

Further performance improvements were achieved through the combined use of lazy loading and asynchronous processing. In low-resource hospital environments, where computing power and memory are often constrained, lazy loading proved particularly effective by deferring non-essential components until explicitly needed. This approach reduced memory overhead and shortened time-to-first-interaction by approximately 50%. Previous studies have demonstrated similar benefits that deferred loading strategies improve responsiveness in data-intensive applications. Jain (2022), for example, reported that combining lazy loading with code splitting in real-world web applications reduced page load times by up to 40% while improving key performance metrics such as First Contentful Paint, Largest Contentful Paint, and Time to Interactive (31). Similarly, Ravalji et al. (2024) demonstrated that optimizing Angular-based dashboards for real-time data analysis through lazy loading, among other techniques can maintain low latency and fast rendering even when handling large, continuously updating datasets (32). Together, these studies underscore the importance of deferred loading and efficient client-side processing for improving responsiveness, scalability, and user experience in data-intensive, real-time dashboard applications.

Complementing these client-side strategies, asynchronous execution of long-running server-side tasks further enhanced interactivity. By offloading computationally intensive processes such as bulk data ingestion and preprocessing across hundreds of facilities, to background execution, user interface rendering improved by nearly 45%, while maintaining responsiveness to user input. This design follows best practices in Shiny development, where non-blocking execution using futures and promises is widely recommended to reduce perceived latency and support scalability in complex, data-driven applications (33, 34). The combined effect of asynchronous processing and lazy loading ensured that users could continue interacting with the dashboard even as resource-intensive operations were executed in parallel.

Additional performance gains were realized through adaptive JavaScript interface optimization, which specifically addressed device-related performance variability. By tailoring JavaScript execution to device capabilities, rendering delays on low-powered hardware were reduced by approximately 30%. This enhancement expanded dashboard accessibility across smartphones and tablets; devices that frontline healthcare workers in resource-limited settings frequently rely upon while preserving seamless interaction and consistent functionality across heterogeneous hardware configurations. These observations are supported by prior evidence on JavaScript performance optimization for low-end devices. Chaqfeh et al. (2021) demonstrated that selectively simplifying and removing non-essential JavaScript components using the JSAnalyzer tool reduced page load times by roughly 30% on low-end mobile phones, while preserving interactivity and improving performance scores by more than 88% (35). Such adaptive strategies are therefore critical for promoting equitable access to digital health tools in settings with constrained device capabilities.

Finally, the adoption of vectorized programming substantially improved the efficiency of computationally intensive operations, yielding positive responsiveness of nearly 50% and streamlining analytical workflows. In R-based systems, vectorization allows operations to be executed at compiled speeds by leveraging optimized low-level code, thereby avoiding the overhead associated with explicit looping constructs. Prior studies have shown that replacing slower string-matching routines with vectorized base functions can accelerate filtering tasks in Shiny applications by more than 30-fold (36). The magnitude of improvement observed in the present study is consistent with these reports and further highlights the critical role of vectorized computation in sustaining responsive, scalable dashboards under high data volumes and complex analytical demands.

Beyond gains in speed and interactivity, the optimized NEST-IT dashboard achieved a system uptime of 99.2%, compared with 92.5% at baseline, reflecting marked gains in reliability and operational stability, key metrics in healthcare environments where interruptions can delay clinical decision-making and compromise continuous care monitoring (37, 38). Moreover, the significant reductions in time-to-first-byte and server request processing underscore improvements in system throughput and backend efficiency, aligning with prior evidence that robust backend responsiveness is central to maintaining real-time usability in digital platforms (23).

### 4.1 Strengths and Limitations

A major strength of this study is the use of empirical pre- and post-implementation comparisons, which quantified performance gains across diverse and complex plots, thereby demonstrating generalizability across real-world use cases. The inclusion of multiple complementary optimization strategies also allowed us to evaluate synergistic benefits. However, a limitation is that our evaluation focused primarily on technical performance metrics (rendering speed, latency reduction, and uptime) rather than end-user outcomes such as clinical decision-making efficiency or satisfaction.

### 4.2 Future Research

Future research should evaluate how these optimizations influence clinical utility and adoption; for example, assessing the effect of improved dashboard performance on time-to-decision in neonatal care could link technical enhancements directly to patient outcomes.

### 4.3 Programmatic Implications

These findings highlight that technical optimization is essential, not optional, for dashboards in resource-limited settings. By reducing delays and ensuring accessibility across devices, the optimized NEST-IT dashboard supports timely monitoring, program evaluation, and accountability for neonatal interventions. Ministries of Health and implementing partners can deploy such dashboards confidently, knowing they remain performant under heavy data loads and constrained computational resources.

## 5 Conclusion

Systematic optimization transforms R Shiny dashboards from slow, fragile tools into responsive, scalable platforms capable of supporting real-time decision-making in neonatal care. Strategies such as reactive programming, caching, and asynchronous processing enabled the NEST-IT dashboard to achieve near-instant interactivity, reduce visualization delays below critical usability thresholds, and maintain stability under heavy use. These insights extend beyond neonatal care, providing practical, context-specific guidance for optimizing R Shiny applications across diverse domains. Optimized dashboards like NEST-IT offer a scalable model for strengthening monitoring and evaluation systems, enabling timely, data-driven actions to improve outcomes globally.

## Data Availability

All data produced in the present work are contained in the manuscript

## Statements

### Data Availability Statement

The data supporting the findings of this study, along with code snippets illustrating the optimization strategies, are provided in the Supplementary Materials. Additional requests for access or clarification can be directed to the corresponding author.

### Author Contributions

JT: Conceptualization, methodology, software optimization, data analysis, writing – original draft, writing – review and editing. GJ: Software optimization, data collection, writing – review and editing. JC: Software optimization, writing – review and editing. MO: Conceptualization, writing – review and editing. LM: Conceptualization, software optimization, writing – review and editing. LRH: Conceptualization, writing – review and editing. RR-K: Conceptualization, writing – review and editing. ZMO: Conceptualization, writing – review and editing. CB: Conceptualization, writing – review and editing, supervision. JW: Conceptualization, methodology, writing – review and editing, supervision.

### Conflict of Interest

The authors declared that the research was conducted in the absence of any commercial or financial relationships that could be construed as a potential conflict of interest.

## Funding

This work is funded through the NEST360 Alliance with thanks to John D. and Catherine T. MacArthur Foundation, the Bill & Melinda Gates Foundation, ELMA Philanthropies, The Children’s Investment Fund Foundation UK, The Lemelson Foundation, The Sall Family Foundation, and the Ting Tsung and Wei Fong Chao Foundation under agreements to William Marsh Rice University.

## 6 Supplementary Material

The Supplementary Material accompanying this article includes illustrative code excerpts detailing the optimization strategies employed within the NEST-IT dashboard, along with the datasets generated during the study.

### 6.1 Supplementary Figures

**Figure S1.**
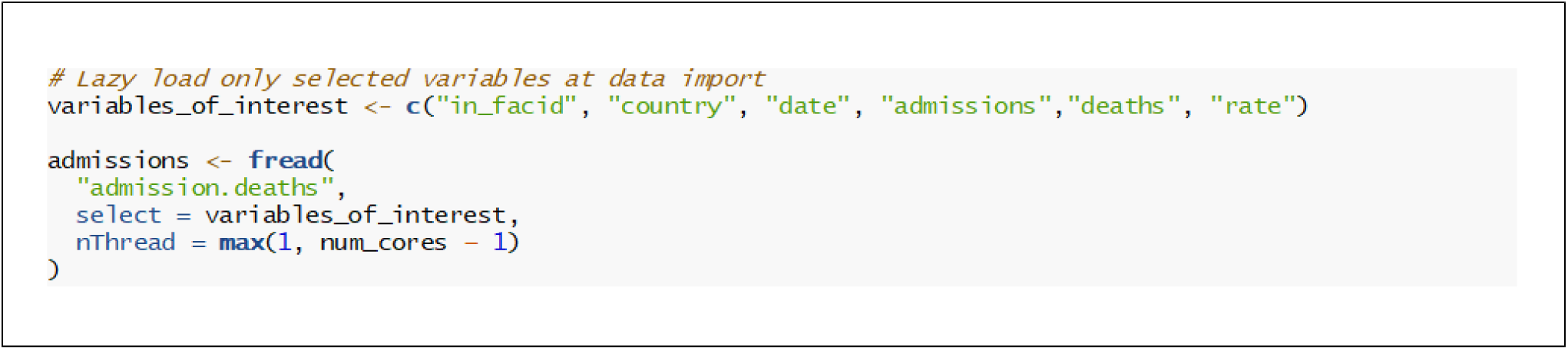
Code snippet demonstrating how to implement lazy loading for efficient data handling *The approach only loads variables explicitly required for a given task into memory, ensuring reduced memory consumption which might create unnecessary computational overhead*

**Figure S2.**
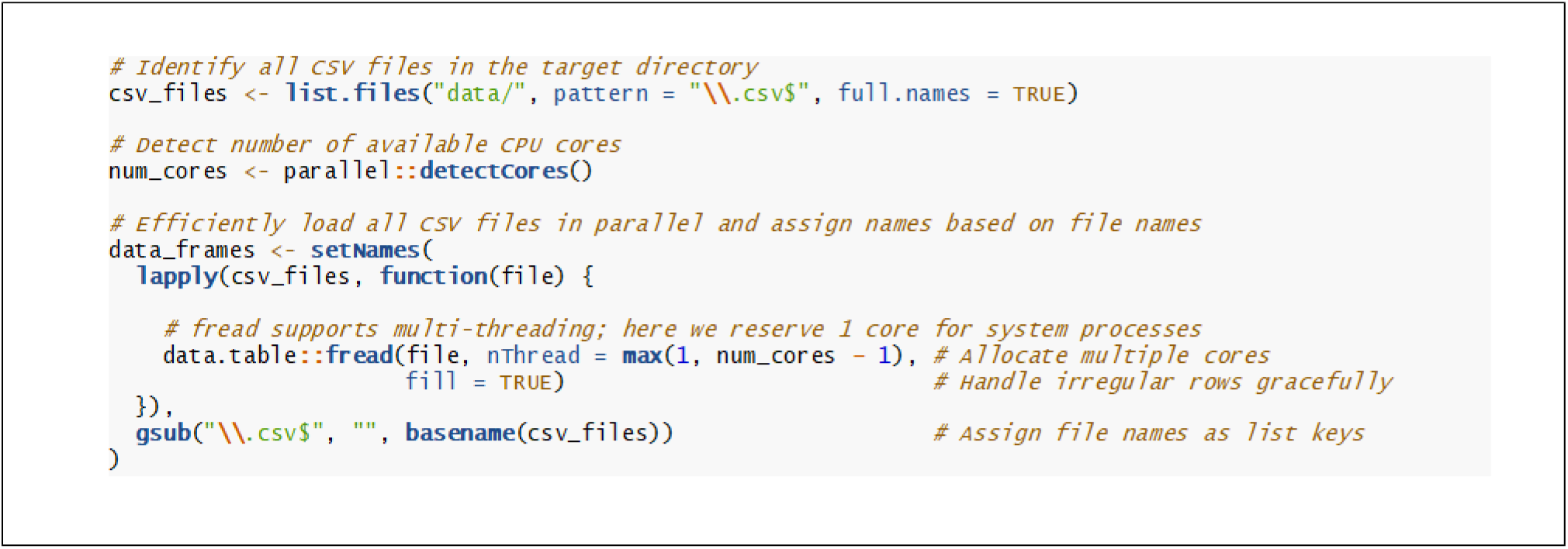
Code snippet demonstrating how to implement parallelized bulk data loading for efficient and reproducible data handling *The fread () function reads each CSV file concurrently, allocating up to num_cores-1 threads to balance performance and system stability. The setNames () function wrapper, automatically assigns human-readable object names based on the source filenames, facilitating organized and reproducible data*

**Figure S3A.**
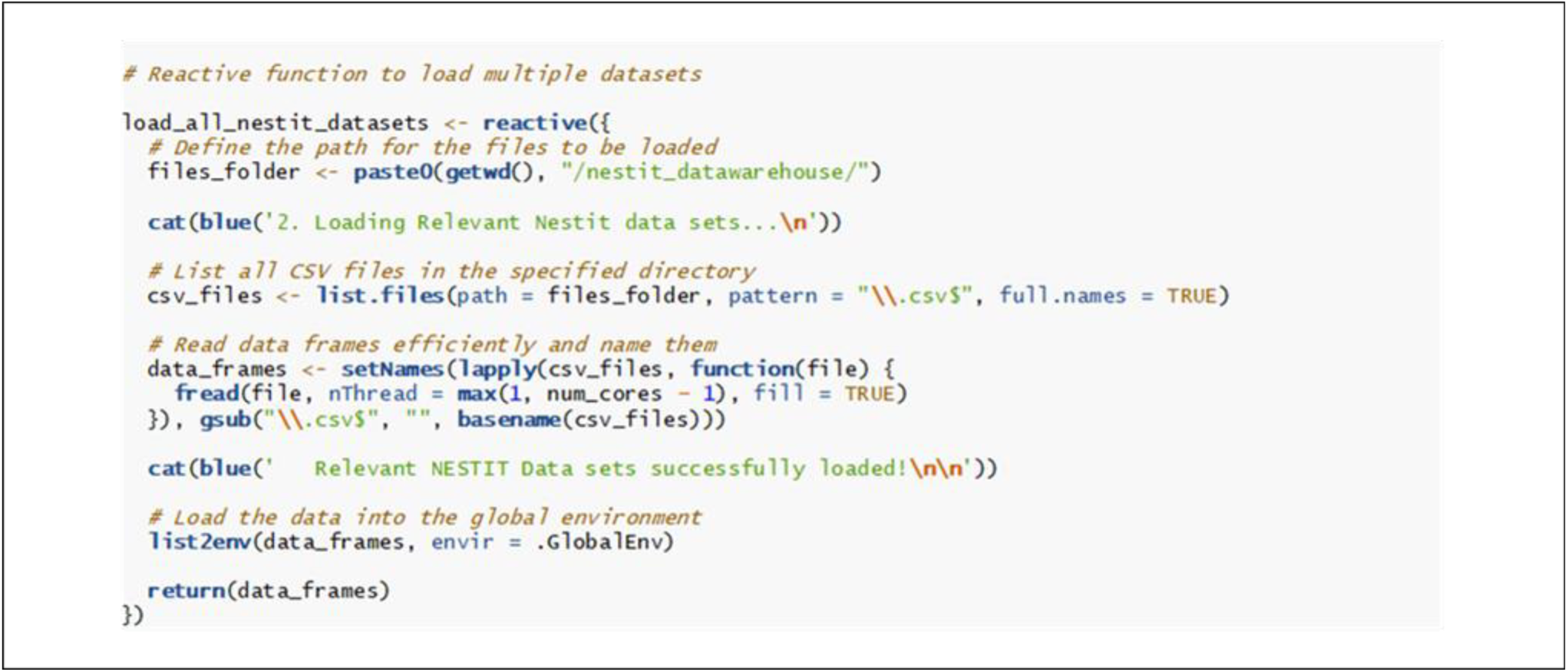
Code snippet demonstrating how to implement reactive programming for timely data update *In this strategy, if any dataset within the list is changed, the reactive data object (reactive ()) detects the update, triggering re-execution of the dependent reactive expressions (load_all_nestit_datasets ()). As a result, the updated dataset automatically refreshes to include the latest numbers*.

**Figure S3B.**
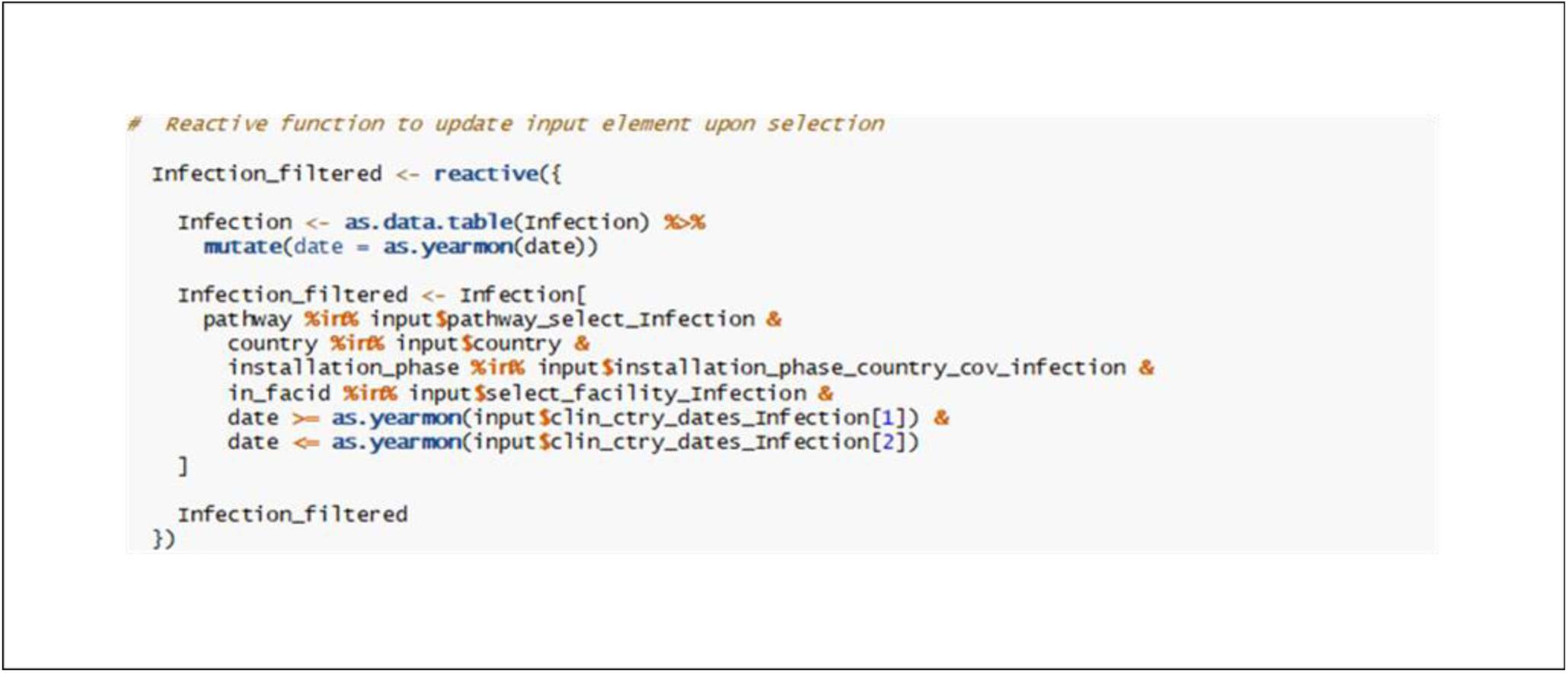
Code snippet demonstrating how to implement reactive programming for timely input element update *When a user modifies the selected date range, the R Shiny framework automatically detects the change in the input $clin_ctry_dates_Infection. The infection_filtered () automatically recomputes updated dataset with the new date range. Any plot or table that uses the infection_filtered () will refresh instantly with the updated data, without reloading the whole data. This reactive structure ensures that only what is necessary is recalculated, making the dashboard fast, responsive, and always synchronized with user selections*.

**Figure S4.**
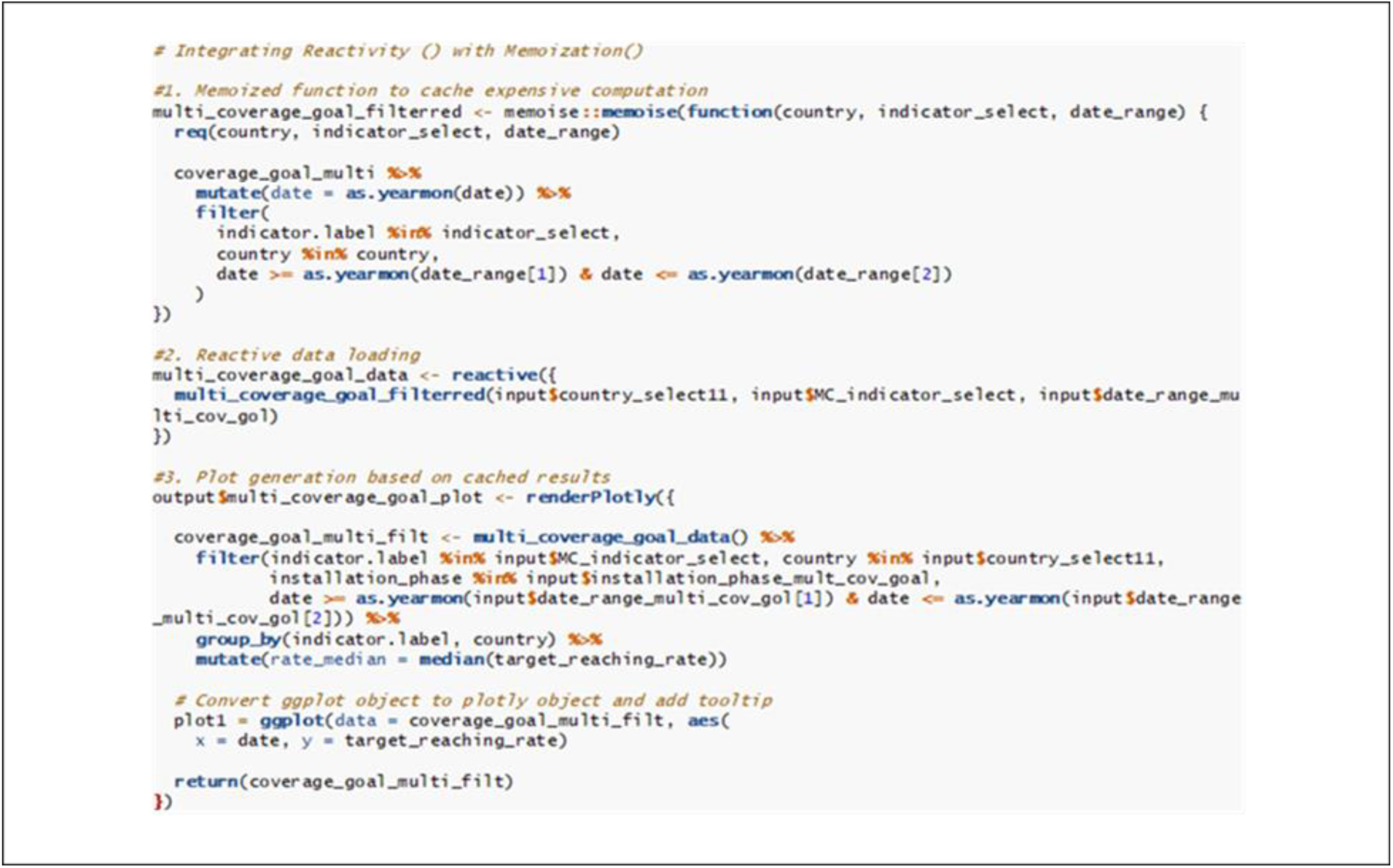
Code snippet demonstrating how to integrate reactive programming with memoization seamlessly for enhanced responsiveness for large, complex dataset *In this implementation, a memoized function first does the heavy lifting by filtering a large dataset (coverage_goal_multi) based on user inputs. By wrapping the function with memoise∷memoise (), repeated requests for the same (country, indicator_select, date_range) combination are served directly from memory cache, bypassing redundant computation. This is especially useful in NEST-IT, where users frequently navigate back and forth between the same filters.* *The reactive data pipeline ensures that whenever the user changes country, indicator or date_range, it automatically triggers the memoized function with the new arguments. If the requested combination has been computed previously, results are retrieved instantly from cache; if not, the function runs once, stores the output, and reuses it in subsequent calls.* *Plots are then generated from these cached subsets rather than re-filtering the full coverage_goal_multi dataset each time. This design significantly accelerates rendering and enhances user responsiveness, which is critical when working with large, multi-country coverage dataset*

**Figure S5.**
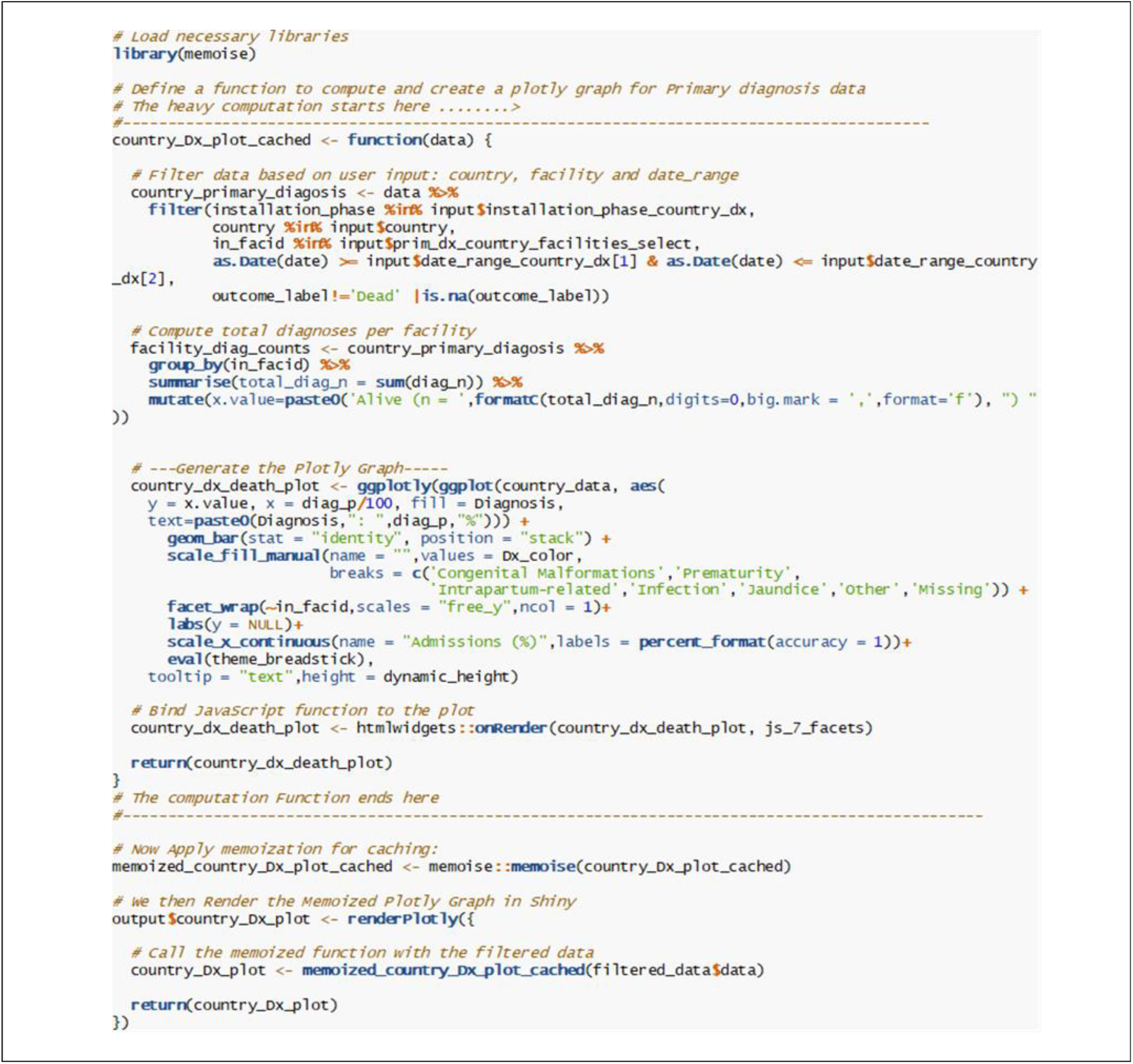
Code snippet demonstrating how to cache computationally expensive computations for enhanced responsiveness *In the NEST-IT dashboard, the function country_Dx_plot_cached () is used to generate a plotly visualization of primary diagnosis data. This function performs several computationally expensive operations, including data filtering, aggregation, and the generation of a faceted stacked bar chart. Without caching, these computations would be repeated every time the user adjusted filters or revisited the same view, leading to longer rendering times. By wrapping the function with memoise (), repeated calls with identical inputs (e.g., same country, facility, and date range) return cached results instantly, bypassing redundant computations*.

**Figure S6.**
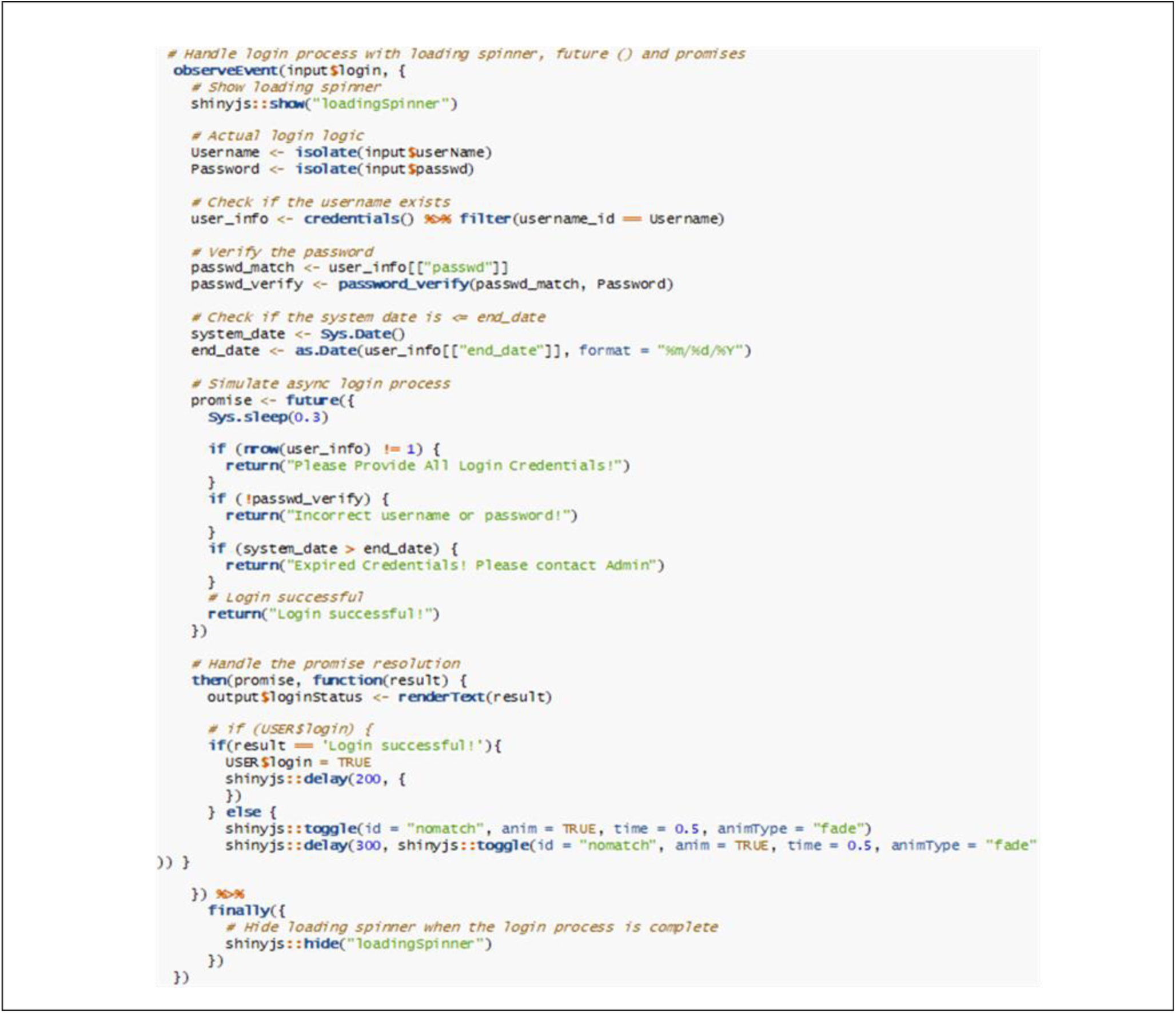
Code snippet showcasing the use of the future () and promises packages to implement asynchronous processing *Asynchronous login handling was implemented using the future () and promises packages to improve system responsiveness during user authentication. This approach allowed credential verification to be executed in the background while simultaneously providing immediate visual feedback to users through a loading spinner. By decoupling the authentication process from the main application thread, the system prevented blocking when multiple users attempted to log in concurrently, thereby supporting smoother and more scalable login performance under real-world operating conditions*.

**Figure S7A.**
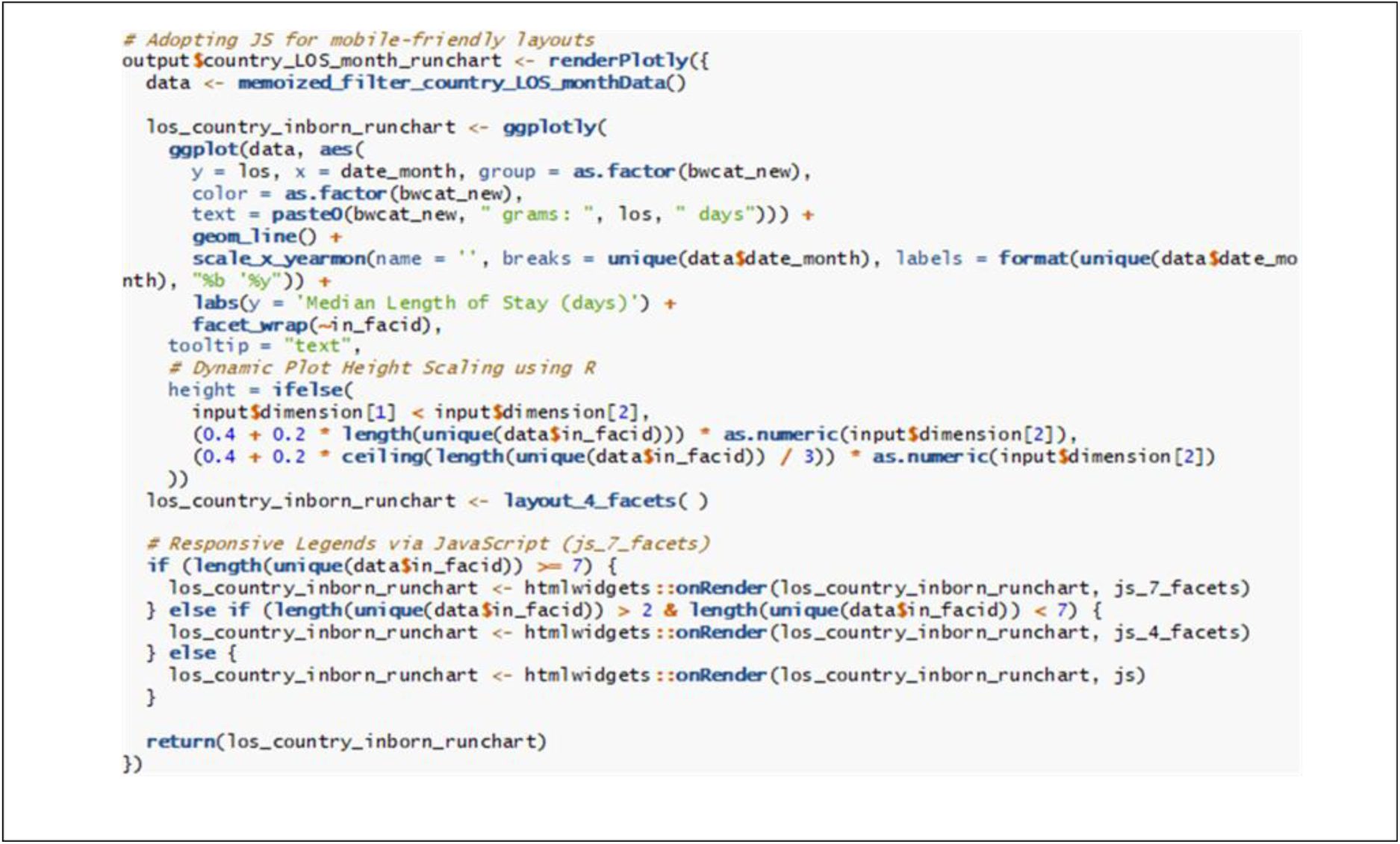
Code snippet illustrating the use of JavaScript for dynamic plot visualization *To prevent vertical compression of plots and maintain readability on smaller screens such as tablets and smartphones, a dynamic plot height scaling mechanism was implemented. This approach automatically adjusts the plot height based on the device’s screen orientation and the number of displayed facets, thereby preserving visualization clarity across different screen sizes*

**Figure S7B.**
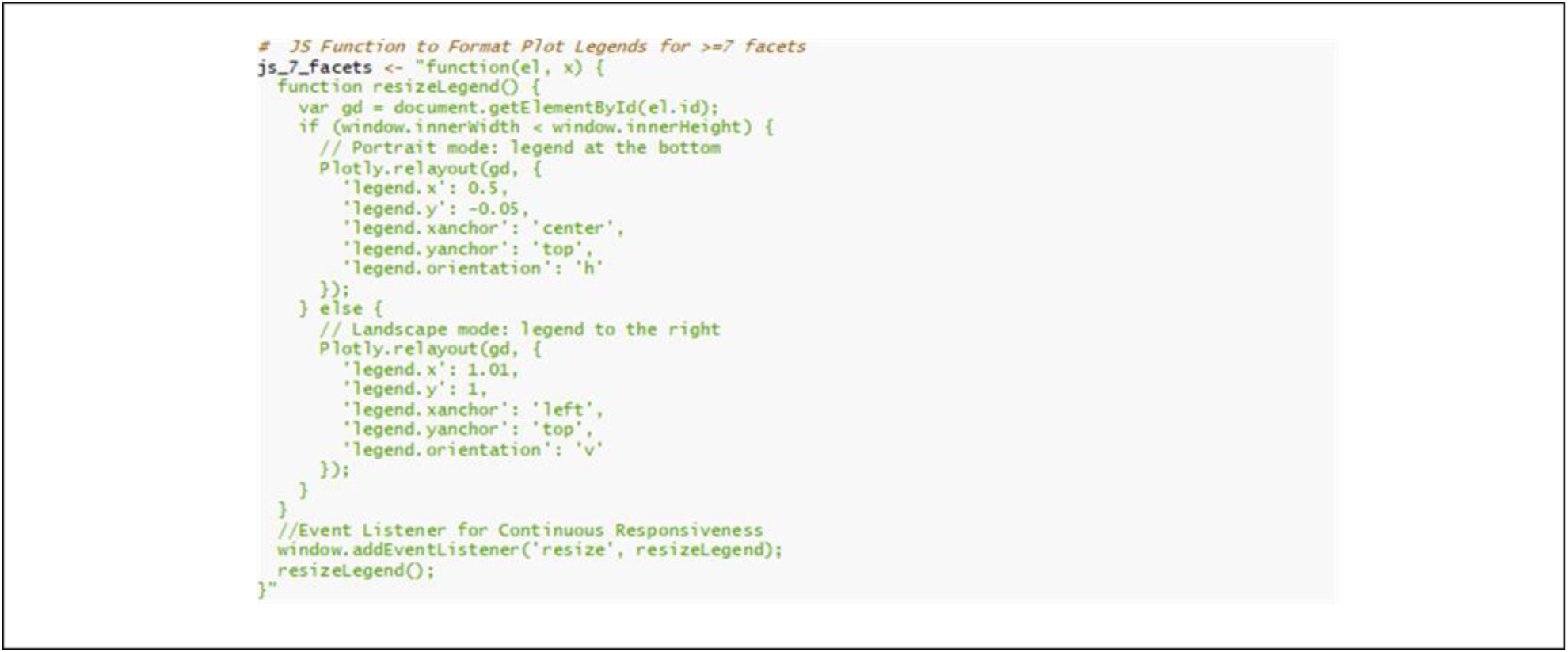
Code snippet demonstrating the use of JavaScript for scaling multi-faceted plots *Also, depending on the number of facets (subplots), different JavaScript scaling logic (js, js_4_facets, js_7_facets) for simple to complex plots, were injected to ensure that even complex multi-country or multi-facility plots remain readable and responsive across devices without overwhelming limited screen state.* *In the above example, the JavaScript scaling logic was implemented for multi-panel plots with ≥7 facets. The legend automatically repositions according to device orientation, displayed horizontally below the plot in portrait mode or vertically to the right in landscape mode. This prevents the legend from encroaching on the data display area, a frequent issue on smaller screens. An event listener further ensures that legend placement updates dynamically in response to screen resizing events*.

**Figure S8.**
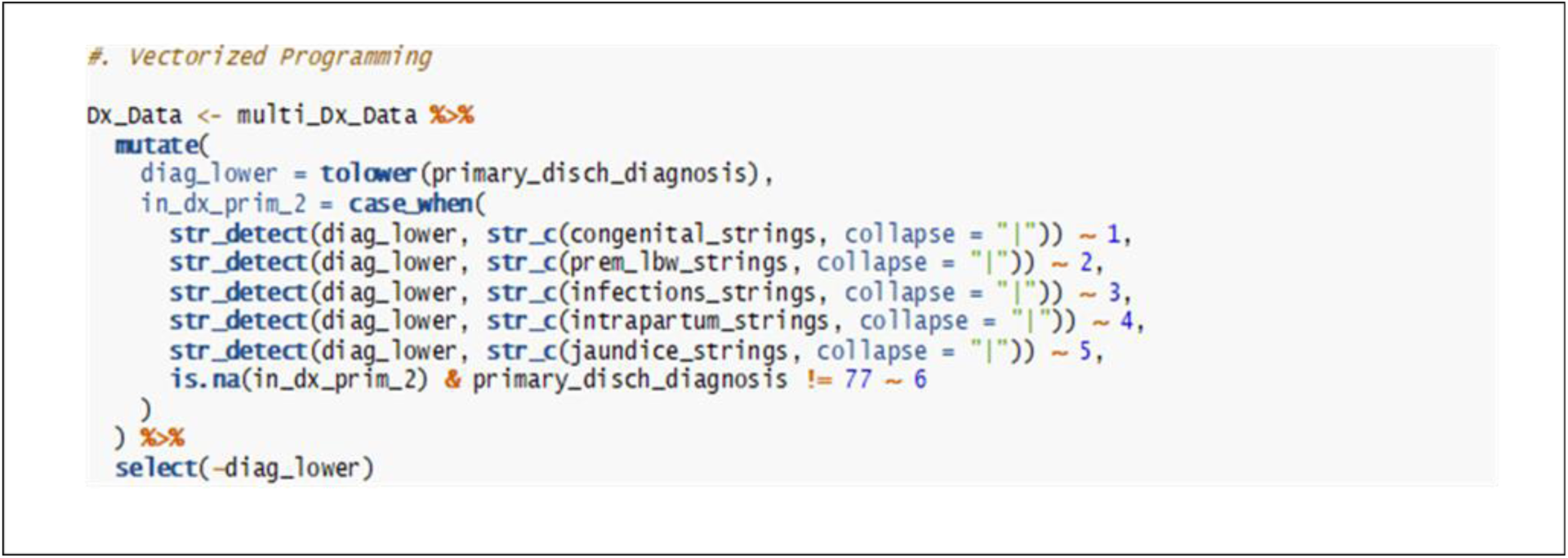
Code snippet demonstrating illustrating the use of vectorized programming to replace computationally intensive iterative operations in large datasets *Rather than using an iterative if–else approach that processes data row by row and becomes computationally expensive for large datasets, we implemented vectorized operations. This strategy reduces redundant transformations, resulting in substantially faster execution while also producing cleaner and more maintainable code*.

### 6.2 Supplementary Data

**Data S1. Visualization Rendering Time Dataset**

This dataset (*XLSX* format) reports the average visualization rendering time (in seconds) measured before and after the implementation of system optimization strategies. The variables capture indicator plot loading times across different sections and hierarchical levels of the NEST-IT dashboard, as well as the mean rendering latency for visualizations across multiple dashboard levels. These measurements enable a comparative assessment of visualization performance and responsiveness following optimization.

**Data S2. Dashboard Performance Metrics Dataset**

This dataset (XLSX format) reports three performance metrics measured during baseline assessment and after implementation of optimization strategies in the NEST-IT dashboard profiling stage. The variables include repeated measurements for each metric and their corresponding average values for comparative analysis

**Data S3. Dashboard Optimization Performance Dataset**

This dataset (XLSX format) reports the optimization performance of the NEST-IT dashboard following the implementation of multiple optimization strategies. It includes mean baseline times, mean times after optimization, and calculated time improvements (%) for each strategy, enabling a quantitative assessment of performance gains. The metrics captured reflect key aspects of system responsiveness, including data processing, plot rendering, and overall.

## Notes

### Competing Interest Statement

The authors have declared no competing interest.

